# Generation and assessment of cytokine-induced killer cells for the treatment of colorectal cancer liver metastases

**DOI:** 10.1101/2023.09.17.23295597

**Authors:** Celine Man Ying Li, Yoko Tomita, Bimala Dhakal, Teresa Tin, Runhao Li, Josephine A Wright, Laura Vrbanac, Susan L Woods, Paul Drew, Timothy Price, Eric Smith, Guy J Maddern, Kevin Fenix

## Abstract

Colorectal cancer (CRC) is the second leading cause of cancer-related death worldwide. Cytokine-induced killer (CIK) cells are an adoptive immunotherapy reported to have strong anti-tumour activity across a range of cancers. They are a heterogeneous mix of lymphoid cells generated by culturing human peripheral blood mononuclear cells (PBMCs) with cytokines and monoclonal antibodies *in vitro*. We compared CIK cells from healthy donors and donors with CRC liver metastases, generated in RPMI supplemented with 10% fetal bovine serum (FBS) or three different serum-free media. CIK cells generated in serum free medium X-VIVO 15 were comparable to those from RPMI medium with 10% FBS in terms of the number and percentages of the main subsets of cells in the CIK culture, and the intracellular levels of granzyme B and perforin, and the pro-inflammatory cytokines IL-2, IFN-γ and TNF-α. There was no significant difference in cytotoxicity against CRC cell lines grown in 2D cultures or as spheroids, and against autologous patient-derived tumour organoids. CIK cells from patients with CRC liver metastases grown in X-VIVO 15 media were similar to those from healthy donors in each of the measures. Donor attributes such as age, sex, or prior chemotherapy exposure had no significant impact on CIK cell numbers or function. These results support further investigations into the therapeutic application of autologous CIK cells in the management of patients with CRC liver metastases.

## Introduction

Colorectal cancer (CRC) is the second leading cause of cancer-related death worldwide [1]. The 5-year survival rate of CRC is 75%, but decreases to 30% with metastatic disease [2]. Currently, 25-70% of patients develop colorectal cancer liver metastases (CRLM) [3, 4]. Treatment options for CRLM patients include surgical resection and chemotherapy, but only a small proportion of CRLM patients are eligible for surgical resection and chemotherapy resistance commonly develops [5]. Thus, there is still a great need for more effective therapies for late-stage CRCs such as CRLM [6].

Cytokine-induced killer (CIK) cell therapy is a cellular adoptive immunotherapy first described in the 1990s [7]. CIK cells are cultured from peripheral blood mononuclear cells (PBMCs) to generate a heterogeneous mix of immune effector cells. Well described subpopulations include conventional T cells (CD3+CD56-), natural killer (NK)-like T cells (CD3+CD56+), and NK cells (CD3-CD56+). The expression of both T cell and NK cell receptors by CIK cells allows for both major histocompatibility complex (MHC)-dependent and independent tumour recognition. CIK cells have been reported to have strong anti-tumour activity across a range of cancers, including solid tumours [8]. CIK cells can induce tumour cytotoxicity by release of cytolytic granules and expression of death ligands (FASL and TRAIL). They also mount a type-1 inflammatory response by releasing interleukin (IL)-2, interferon (IFN)-γ and tumour necrosis factor (TNF)-α [7, 9]. Meta-analyses of CRC clinical trials for CIK cell therapy, mostly conducted in China, have shown significant improvement in patient outcomes, including overall and progression-free survival [10, 11].

An advantage of CIK cell therapy is its relative ease of production and inexpensive material costs. Briefly, PBMCs are cultured for up to 28 days in the presence of IFN-γ, anti-CD3, and IL-2 [8]. Quality control of the culture product is currently based on counting viable CD3+CD56- and CD3+CD56+ cells by flow cytometry. It is generally considered that CIK cells for patient infusion should contain about 90% CD3+ T cells with expansion of T cells co-expressing CD56. While there is broad consensus on how to generate CIK cells for laboratory studies, there is not a universally agreed clinical standard operating procedure for the generation of CIK cells for clinical use. Most laboratory and some clinical studies generated CIK cells in media supplemented with fetal bovine serum (FBS) [12-15]. However, animal-derived serum components have the potential to increase the risk of adverse events [16] and thus are not suitable for use in clinical good manufacturing practice (GMP) protocols. There are now a wide variety of GMP suitable serum free media (SFM) available for T cell therapy production that may be suitable for CIK cell cultures.

In this study we identified X-VIVO 15 as a suitable GMP compliant T cell culture specific SFM to expand CIK cells from PBMC of healthy and CRLM donors. We assessed CIK cell numbers and functionality including cytotoxicity to tumour cells. We examined how patient characteristics such as age, sex, and prior chemotherapy exposure affected CIK cell production and functionality. This study supports the application of autologous CIK cell therapy as a potential treatment for patients with CRLM.

## Materials and Methods

### Study Group

Healthy donors or patients with CRLM were consented at The Queen Elizabeth Hospital (TQEH, Woodville, South Australia). Donor data are recorded in Table 1 and Supplementary Tables 1 and 2. Donor Patient IDs were generated by the research group and cannot be used to identify patients external to the group. This study was approved by the Human Research Ethics Committee of the Central Adelaide Local Health Network under protocol number HREC/14/TQEHLMH/164.

**Table 1.**
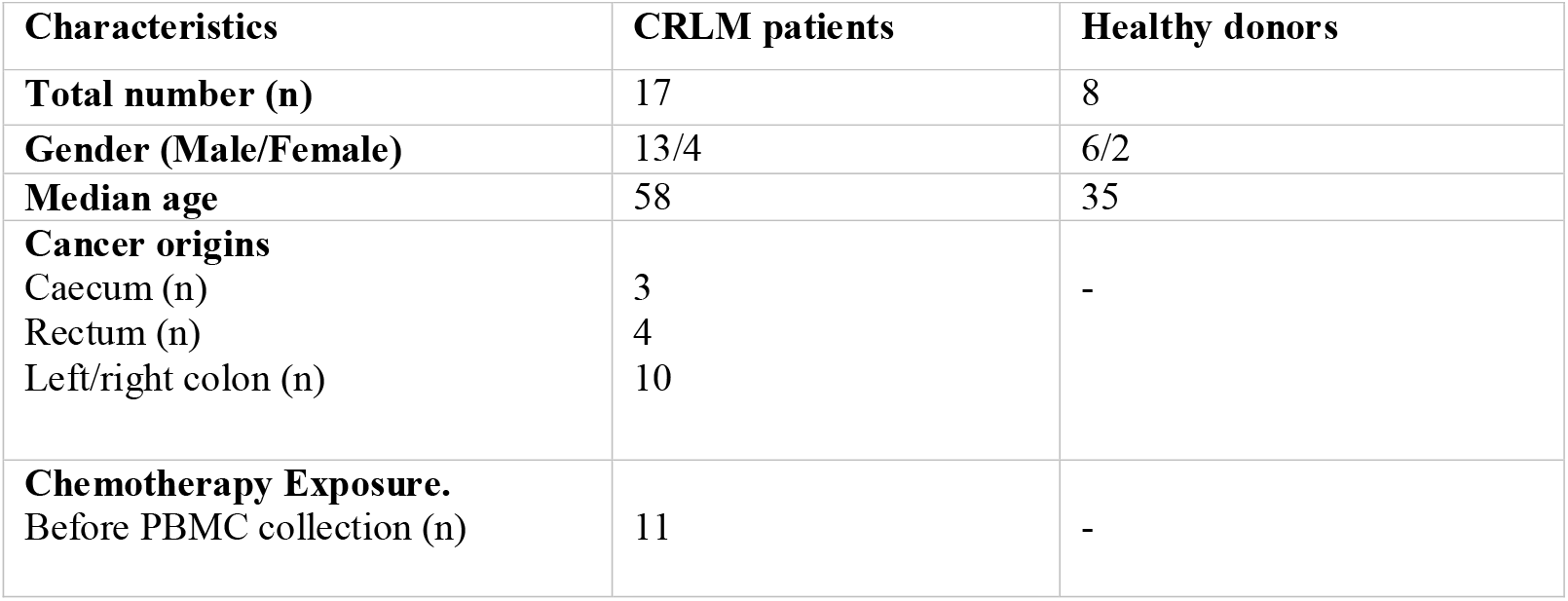
Characteristics of healthy donors and patients with liver colorectal cancer metastases.

| Characteristics | CRLM patients | Healthy donors |
| --- | --- | --- |
| **Total number (n)** | 17 | 8 |
| **Gender (Male/Female)** | 13/4 | 6/2 |
| **Median age** | 58 | 35 |
| **Cancer origins** |  |  |
| Caecum (n) | 3 | - |
| Rectum (n) | 4 |  |
| Left/right colon (n) | 10 |  |
| **Chemotherapy Exposure.** |  |  |
| Before PBMC collection (n) | 11 | - |

### CIK Cell Generation

The PBMCs were isolated from donor venous blood using Ficoll-Paque (Bio-Strategy, USA) density gradient centrifugation as previously described [17]. The patient blood was collected just before curative intent liver resection for CRLM. Cells were cultured in a 24- or 12-well tissue culture grade plates. Complete RPMI is defined as RPMI 1640 medium (Life Technologies, USA) containing 10% FBS (Sigma-Aldrich, USA), 1X L-Glutamine (Gibco, USA), and 200 U/mL penicillin and 200 μg/mL streptomycin (pen-strep) (Life Technologies, USA). Serum-free media, X-VIVO 15 (LONZA, Switzerland), TexMACS (Milltenyi Biotec, USA), and CTS OpTimizer (Gibco, USA) were supplemented with pen-strep. Briefly, isolated PBMCs were seeded at 2 × 10^6^ cells/mL in media containing 1000 U/mL of interferon gamma (IFN-γ) (Miltenyi Biotec, USA). After 24 hours, 0.05 μg/mL of anti-CD3 (Miltenyi Biotec, USA) and 300 IU of interleukin (IL)-2 (Miltenyi Biotec, USA) were added. Three days post anti-CD3 stimulation, culture media were topped up with 500 μL of media containing 300 IU of IL-2. The cells were split every 3-4 days and seeded at density of 1 × 10^6^ cells/mL with the addition of 300 IU IL-2 for up to 21 days from isolation of the PBMCs.

### Flow Cytometry

Cells were stained with BD Horizon Fixable Viability Stain 780 (FVS780) (Biolegend, USA). Cells were treated with 50 μL of Fc block (BD Biosciences, USA), and then stained with anti-human CD3 V510, anti-human CD56 PeCy-7, anti-human CD4 FITC, anti-human CD8 647, anti-human CD226 PE, and anti-human CD314 V421 (Biolegend, USA) prepared in FACS buffer (2% FBS, 0.05% sodium azide and 1mM EDTA into sterile 1X PBS) for 30 mins at 4°C. After washing with FACS buffer, the fluorescence data was acquired using a FACS Canto II flow cytometer (BD Biosciences, USA). For measurement of intracellular markers, cells were stimulated and cultured with 1X Cytokine Activation Cocktail (BD Biosciences, USA) for 5 hours. They were then permeabilised with Intracellular Fixation Buffer (BD Biosciences, USA) for 20 mins and washed with permeabilization buffer/wash buffer (BD Biosciences, USA). They were stained with anti-human granzyme B BV421 and anti-human perforin PerCP5.5 for intracellular cytotoxicity markers or with anti-human tumour-necrosis factor (TNF)-α PerCP5.5 and anti-human interferon IFN-γ BV421 and anti-human IL-2 PE for pro-inflammatory cytokines. Cells were resuspended in 150-200 μL of FACS buffer and the fluorescence data was acquired in the FACS Canto II Flow Cytometry system. The data were analysed by FlowJo v10.8.1 software (BD Biosciences, USA).

### Cryopreservation

The CIK cells were centrifuged and resuspended in FBS with 10% DMSO at a concentration of 1x10^6^ cells/mL. Aliquots in cryovials were frozen using a Mr. Frosty Freezing Container (Thermo Fisher Scientific, USA) at - 80°C overnight before being stored long term in liquid nitrogen. To thaw out the cells, the vial was defrosted in a 37 °C water bath. Immediately after thawing, the cells were gently transferred in a drop-wise fashion into a 15 mL falcon tube containing 1 mL pre-warmed culture media.

### Cell Lines

The CRC cell lines (HT-29, SW620, SW480, COLO 205) were obtained from the American Type Culture Collection (ATCC, USA). HT29 and COLO 205 were maintained in RPMI supplemented with 10% heat-inactivated FBS, 200 U/mL penicillin, 200 μg/mL streptomycin and 200 mM GlutaMAX Supplement (Life Technologies, USA). SW620 and SW480 were maintained in DMEM (Life Technologies, USA) with the same supplementation as RPMI. Cells were incubated at 37 °C with 5% CO_2_.

### Patient-Derived Tumour Organoids

To establish patient-derived tumour organoids (PDTOs), tumour samples were collected from patients undergoing liver resection for CRLM disease and cultured as described previously [18]. Briefly, CRLM tissues were minced and digested in organoid digestion buffer containing 2.5% of FBS, 75 U/ml Collagenase Type IV (Gibco, USA), 125 μg/ml dispase (Gibco, USA), 20 μg/mL hyaluronidase (Sigma Aldrich, USA) and 10 μM Y27632 (Sigma-Aldrich, USA) in advanced DMEM media (Gibco, USA) for 30-60 mins in a water bath at 37 °C. After the tissues were completely digested, single cells were obtained and pelleted by centrifugation. If pellets were contaminated with red blood cells, red blood cell lysis was performed using ACK lysis buffer (Gibco, USA). The pellet was resuspended in a volume of pre-thawed phenol-red free Matrigel (Life Technologies, USA) at 4 °C depending on the density of the cells. Single cells were embedded in the 50 μL Matrigel domes and were cultured in 5-6% low oxygen conditions at 37 °C incubator for 30 mins. Matrigel domes were then topped with CRC media containing advanced DMEM media, 10 mM HEPES, 1X GlutaMAX Supplement, 1X antibiotic-antimycotic, 10 mg/L Gentamicin, 2X B27 (all from Life Technologies, USA), with the addition of 500 nM A 83-01 (Tocris Bioscience, Bristol, UK), 50 ng/mL hEGF, 1nM Gastrin, 1 mM N-acetyl-L cyst, 5 μM SB202190, 10 μM SB431542, 10 μM Y27632 (all from Sigma Aldrich, USA). Organoids were maintained in the same media at 550 μL, and were passaged every one to two weeks, or when they reached 100-200 mm in diameter, by digestion with TrypLE (Life Technologies, USA) at 37 °C.

### PDTO Cytotoxicity Assay

The PDTO cytotoxicity assay was performed as previously described [19]. PDTOs were cultured in a 24-well plate. First, a single well was harvested for cell number estimation. Briefly, PDTOs were washed with ice-cold 1X PBS twice and digested into single cells using 5 mL of TrypLE supplemented with 10 μM Y27632 at 37 °C for less than 15 mins. Then 50 μL of FBS was added to stop digestion before the cell count was performed. The rest of the PDTOs were then harvested and labelled with a 1 in 4,000 dilution of CellTrace™ Violet (Invitrogen, USA) for 20 mins in the dark at room temperature. The PDTOs were seeded into a 96-well flat bottom plate at an equivalent of 1 × 10^5^ cells per well. CRC media supplemented with 10 μg/mL DNase and 300 IU IL-2 was then added into each well. The CIK cells were resuspended in CRC media and co-cultured with PDTOs at effector to target (E:T) ratios of 10:1, 5:1 or 1:1 for 24 hours at 37°C. After 24 hours of incubation, supernatants were collected and 100 μL of TrypLE were added into each well for single-cell dissociation followed by viability staining. Samples were then acquired using a Cytek Aurora spectral flow cytometer (Cytek Biosciences) with volumetric counting. For quantification of the percentage of specific lysis, the following formula was used:

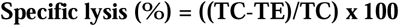

where TC indicates the cell count of live labelled target cells in the control well (target cells alone) and TE indicates the cell count for live labelled target cells in the treatment well (target cells + effector cells) [20].

### CRC Cell Line Cytotoxicity Assay

HT-29, COLO 205, SW480 and SW620 cells, stained with CellTrace™ Violet, were seeded at 1 × 10^4^ cells per well in 96-well flat bottom plates and incubated for 24 hours at 37°C. Detached non-viable cells were removed by washing with Dulbecco’s phosphate buffered saline (DPBS) (Life Technologies, USA), CIK therapy products at 10:1, 5:1 or 1:1 E:T ratio were then added co-cultured with the remaining attached cells for 24 hours. All cells were then harvested by pooling non-adherent cells and the adherent cells were detached by incubation with trypsin at 37 °C for 4 min. The detached adherent cells were then washed three times with DPBS and pooled with non-adherent cells. Samples were labelled with BD Horizon FVS780 viability dye and measured using a Cytek Aurora spectral flow cytometer. Specific lysis was calculated using the same formula as the PDTO cytotoxicity assay.

### 3D Live Cell Imaging Cytotoxicity Assay

Caspase 3/7 activation in response to CIK cells was measured in HT-29 CRC spheroids [21]. HT-29 cells were seeded in a 96-well round bottom ultra-low attachment plate (Corning, New York, NY, USA) at 1 × 10^5^ cells per well and incubated for 72 hours to form 3D spheroids. The CIK cells were added to the spheroids to a final ratio of 10:1, 5:1 or 1:1 ratio together with 1 μM CellEvent Caspase-3/7 Green Detection Reagent (Thermo Fisher Scientific, Waltham, MA, USA). The activation of caspase 3/7 was monitored for the following 24 hours of incubation and the results were captured and analysed using an Incucyte S3 Live-Cell Analysis System (Sartorius, Germany).

### Statistical Analysis

Statistical analyses performed are described in the figure legends. All data were analysed using GraphPad Prism Version 9 (GraphPad Software, USA).

## Results

### 3.1 Impact of serum-free media (SFM) in the generation and functionality of CIK cells

We first compared CIK cells from the PBMC of healthy donors generated using three GMP-grade SFMs (X-VIVO 15, TexMACS and CTS OpTmizer) or complete RPMI (supplemented with 10% FBS). The only SFM which supported similar expansion of total cells compared to complete RPMI was X-VIVO 15 (Fig. 1a and Supp. Fig. 1). Expansion of the CD3+CD56+ cell subpopulation was observed in all media, however only X-VIVO 15 had numbers and percentages close to those in complete RPMI (Fig. 1b). As expected, CD3+CD56- cells were the largest subpopulation in all media tested. X-VIVO 15 supported expansions of CD3+CD56- cells and CD3+CD56+ cells to 13.36 (2.08-27.42) and 4.54 (0.86-12.7) x 10^6^, respectively (Fig. 1c and Supp. Table 3). Mature CD3+CD56- and CD3+CD56+ populations can be further as CD4+ T helper cells or CD8+ cytotoxic T lymphocytes (CTLs) [22]. CIK cells have been shown to consist mostly of CD8+ cells [23]. CIK cells grown in complete RPMI led to CTL expansion that did not differ from X-VIVO 15 with median of 48.7 (16.6-71.4) and 71.4 (43.3-88.7) x 10^6^, respectively (Fig. 1d-e and Supp. Table 4).

**Figure 1.**
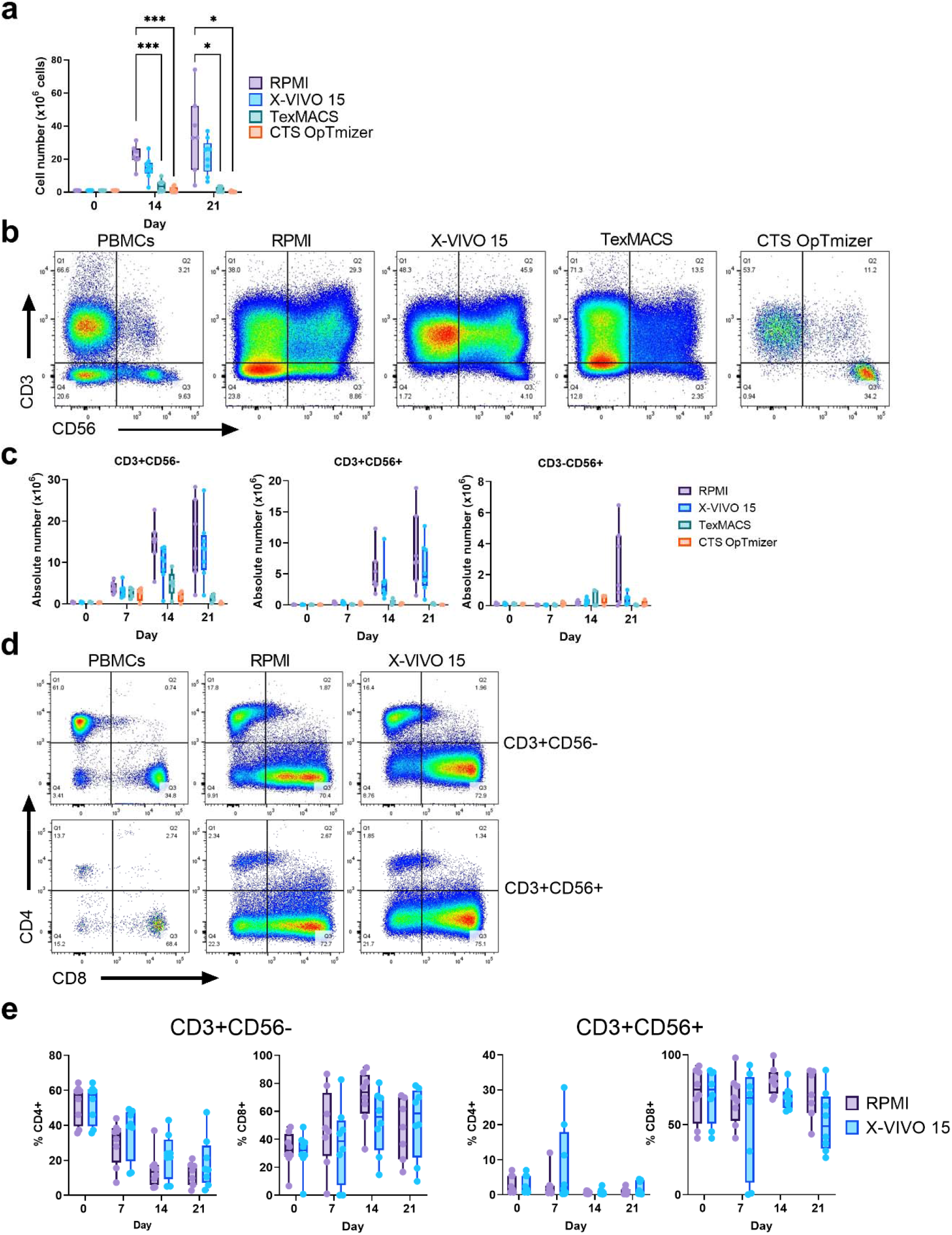
Impact of serum-free media on CIK cell production. CIK cells were generated from healthy donor PBMCs using three serum-free media (X-VIVO 15, TexMACS or CTS OpTmizer) or RPMI supplemented with 10% FBS. (a) Total cell counts on Day 0, 7, 14, and 21 of culture. (b) Representative flow cytometry plots of PBMCs and CIK cells at day 21 post-culture and (c) absolute number counts of T cells (CD3+CD56-), NK-like T cells (CD3+CD56+), NK cells (CD3-CD56+) at different culture time-points. (d) Representative flow cytometry plots showing CD4+ and CD8+ subsets within the CD3+CD56- and CD3+CD56+ subpopulations at day 21 of culture in RPMI or X-VIVO 15 media. (e) Quantitation of CD4 and CD8 single positive subpopulations within CD3+CD56- cells and CD3+CD56+ cells between RPMI and X-VIVO 15 media. Box plots represent the median with each point representing an individual donor. *\*p* ≤ *0*.*05, ***p* ≤ *0*.*005*. Two-way ANOVA with multiple comparisons test were performed to compare the different media.

Next, we determined if CIK cells in X-VIVO 15 had similar capacity against cancer cells as those generated in complete RPMI. First, CIK cells are mostly CD8+CD3+CD56- and CD8+CD3+CD56+ cells, which mediate cell death by the release of cytolytic granules including perforin and granzymes during immunological synapse with their targets [24]. The intracellular expression of granzyme B and perforin in CD8+CD3+CD56+ cells did not differ between CIK cells generated in X-VIVO 15 or RPMI, consistent with these cells having similar cytolytic capacities (Fig. 2a-b). Similar findings were observed for CD8+CD3+CD56- cells (Supp. Fig. 2a).

**Figure 2.**
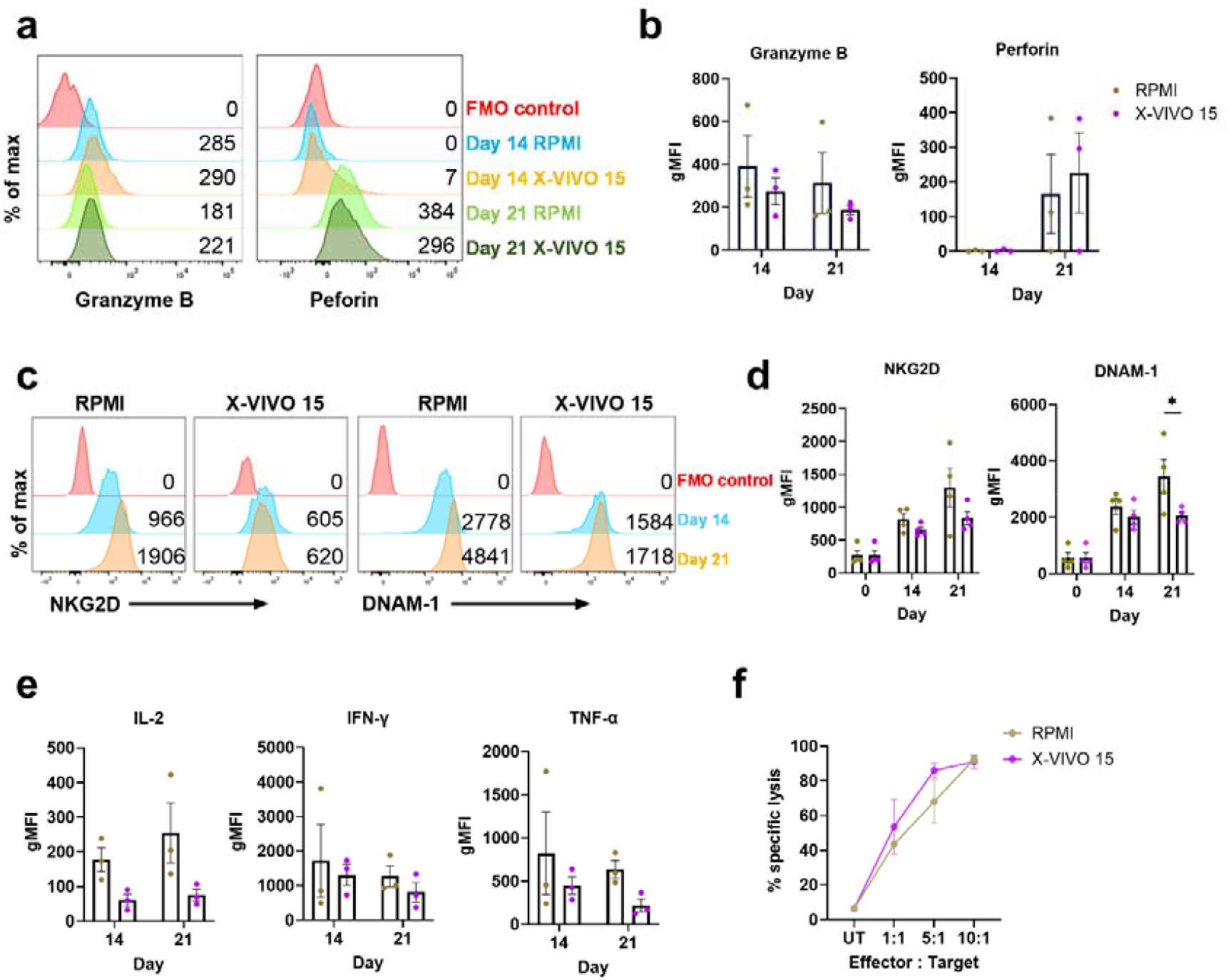
Properties of CIK cells generated in X-VIVO 15 or RPMI. Flow cytometric analysis of CIK cells identified by live CD8+CD3+CD56+ cells. (a) Representative histograms and (b) the geometric mean fluorescence intensity (gMFI) of intracellular granzyme B and perforin (n=3). (c) Representative histograms and (d) gMFI of NKG2D and DNAM-1 expression (n=4). (e) The gMFI for intracellular IL-2, IFN-γ and TNF-α. (n=3). (f) 2D cytotoxicity assay with the HT-29 CRC cell line as target cells and CIK cells at 10:1, 5:1, 1:1 effector to target (E:T) ratios. Target cells only were used as untreated (UT) controls (n=3). *\*p* ≤ *0*.*05*. Two-way ANOVA with multiple comparisons test between the mean of RPMI and X-VIVO 15 media.

CIK cells can recognise cancer cells by MHC-independent mechanisms through NK cell receptors [23]. Expression of NKG2D and DNAM-1, the main NK cell-activating receptors that lead to granule exocytosis, cytokine secretion and cellular cytotoxicity in CIK cells [25]. Expression of both receptors in the CD8+CD3+CD56+ cells increased significantly during culture (Fig. 2c-d). Further, the expression of NKG2D and DNAM-1 were similar in both CD4+ and CD8+ subpopulations of the CD3+ cells and the CD3+CD56+ cells grown in the two media. However, DNAM-1 expression was greater in CIK cells grown in RPMI compared to those grown in X-VIVO 15 at day-21 post-culture (Fig. 2d) (Supp. Fig. 2b).

We then determined if CIK cells grown in X-VIVO 15 produced IL-2, IFN-γ and TNF-α, type-1 cytokines that are commonly associated with CTLs and are expressed by CIK cells [26]. Flow cytometric intracellular staining for these cytokines in the CD3+CD56+ cells, showed less expression in cells grown in X-VIVO 15 than RPMI (Fig. 2e and Supp. Fig 3). Finally, we examined the cytotoxic capacity of these cells against HT-29 CRC cell lines grown as 2D monolayers. There was a dose specific cytotoxic response against target cells, with no difference observed between cells grown in X-VIVO 15 or RPMI (Fig. 2f).

Recently it was reported that CIK cells can be cryopreserved with no significant loss of *in vitro* or *in vivo* cytotoxic potency [27]. Since treatment regimens for CIK therapy include multiple rounds of CIK infusion over many months [5, 28], the ability to be able to use frozen aliquots from one large batch of CIK cells might be of practical benefit. We thus investigated if cryopreservation storage time affects the cytotoxic capacity of CIK cells. Comparing cells stored in liquid nitrogen either for 1-4 or 6-12 months showed that the longer-term cryopreservation can significantly reduce cytotoxic activity (Supp. Fig. 4), suggesting that cryopreservation, while useful, should be for shorter term storage. Together, these data suggest that X-VIVO 15 is a suitable SFM for the generation of CIK cells.

### 3.2 Characterisation of CIK cells derived from CRLM donors

We recently reported that past clinical studies on CIK cell therapy for CRC were generated from autologous patient-derived PBMCs [11]. Thus, we investigated if PBMCs from CRLM donors have similar capacity to produce CIK cells as healthy donors. In total, 16 CRLM donors were recruited for this study (Table. 1). PBMCs from both CRLM and healthy donors could generate CIK cells, although the number of cells expanded varied greatly, with some PBMCs from both healthy and CRLM donors failing to expand (Fig. 3a). In the samples that successfully expanded, the numbers and percentages of CD3-CD56+, CD3+CD56-, and CD3+CD56+ cells did not differ significantly between CRLM and healthy donors (Fig. 3b-d). The percentages of CD4+ and CD8+ subpopulations in the CD3+CD56- and CD3+CD56+ cells were not significantly different between the donor groups. As expected, there were significantly more CD8+ than CD4+ cells (Fig. 3e). We compared the functional capacity of the CD3+CD56- and CD3+CD56+ subpopulations of patients and controls and found similar expression of cytolytic molecules: granzyme B and perforin (Fig. 3f) and type-1 cytokines, IL-2, IFN-γ and TNF-α (Fig. 3g). These results indicate that PBMC from donors with CRLM can produce CIK cells in equivalent numbers and with equivalent expression of functional molecules, consistent with the possibility of them being functionally equivalent.

**Figure 3.**
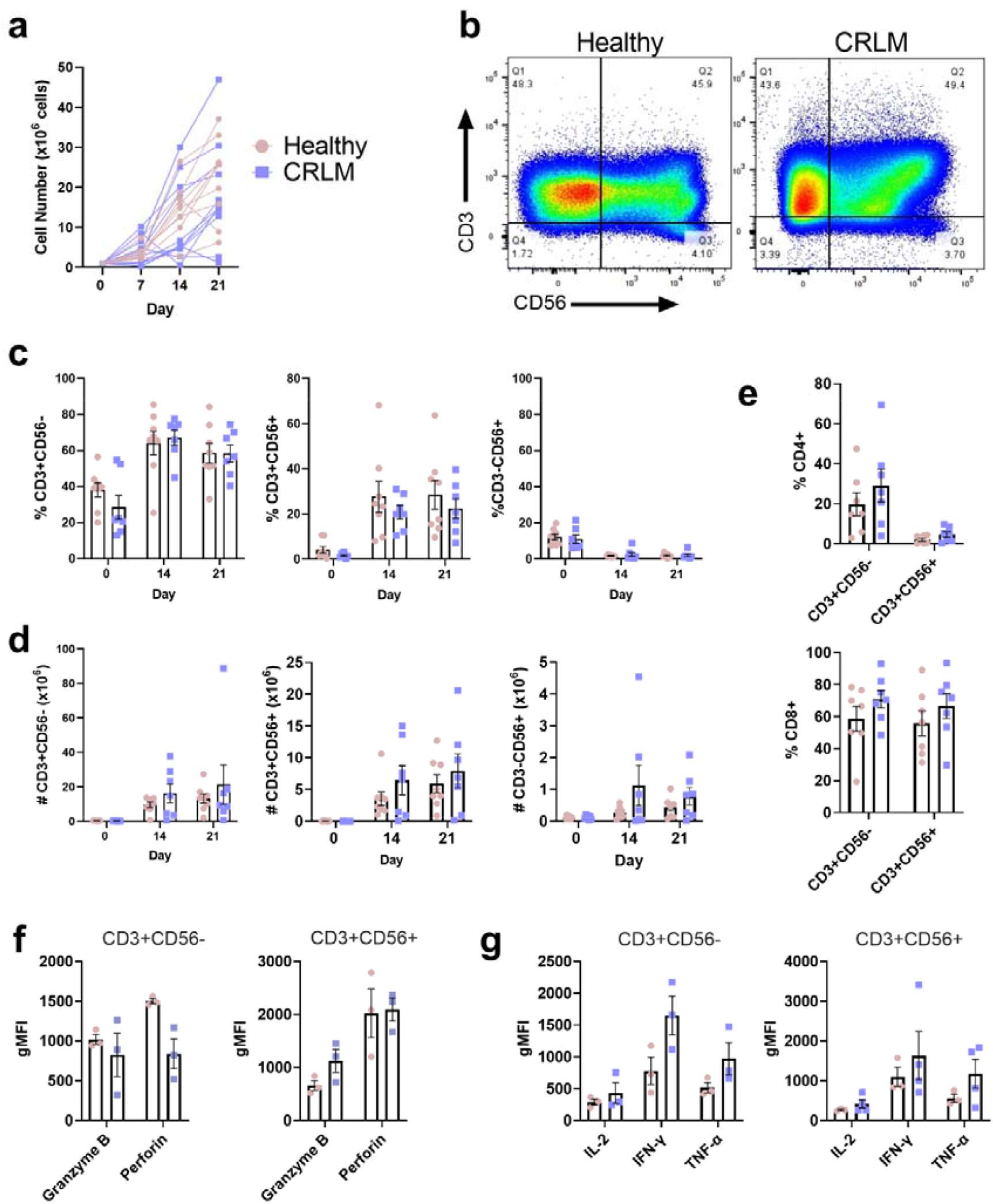
Comparison of CIK cells from CRLM and healthy donors. CIK cell expansion of PBMC from CRLM or healthy donors grown in X-VIVO 15 media. (a) Total cell counts on days 0, 7, 14, and 21 of culture. (b) Representative flow cytometry plots in PBMCs and CIK cells at day 21 of culture and (c) percentage and (d) absolute number counts of T cells (CD3+CD56-), NK-like T cells (CD3+CD56+), NK cells (CD3-CD56+) at different cell culture time points. (e) The percentage of CD4+ and CD8+ subsets within the CD3+CD56- and CD3+CD56+ subpopulations at day 21 of culture. (f) The gMFI for intracellular granzyme B and perforin and (g) IL-2, IFN-γ and TNF-α in CD8+CD3+CD56- and CD8+CD3+CD56+ CIK cells. Data are shown as mean ± SEM with each point representing an individual donor.

Next, we compared the cytotoxic activity of CIK cells from CRLM and healthy donors using several cytotoxicity assays. In 2D cultures using CRC cell lines HT29, COLO 205, SW480 and SW620, there was comparable dose-dependent cytotoxicity between healthy and CRLM-derived CIK cells (Fig. 4a). There was also no difference between CIK cells in the induction of cell death in HT-29 spheroids (Fig. 4b-c). PDTOs are currently considered the best preclinical model to predict patient response to anti-cancer drugs and immunotherapy, including adoptive cell therapies [29, 30]. Using a well-described tumour organoid cytotoxicity assay [19], we confirmed that our CRLM donor-derived CIK cells can eliminate matched PDTOs (Fig. 4d-e). Together, these data indicate CIK cells generated from CRLM donors do not differ in *in vitro* cytotoxic capacity to CIK cells prepared from healthy donors and are able to kill tumour organoids.

**Figure 4.**
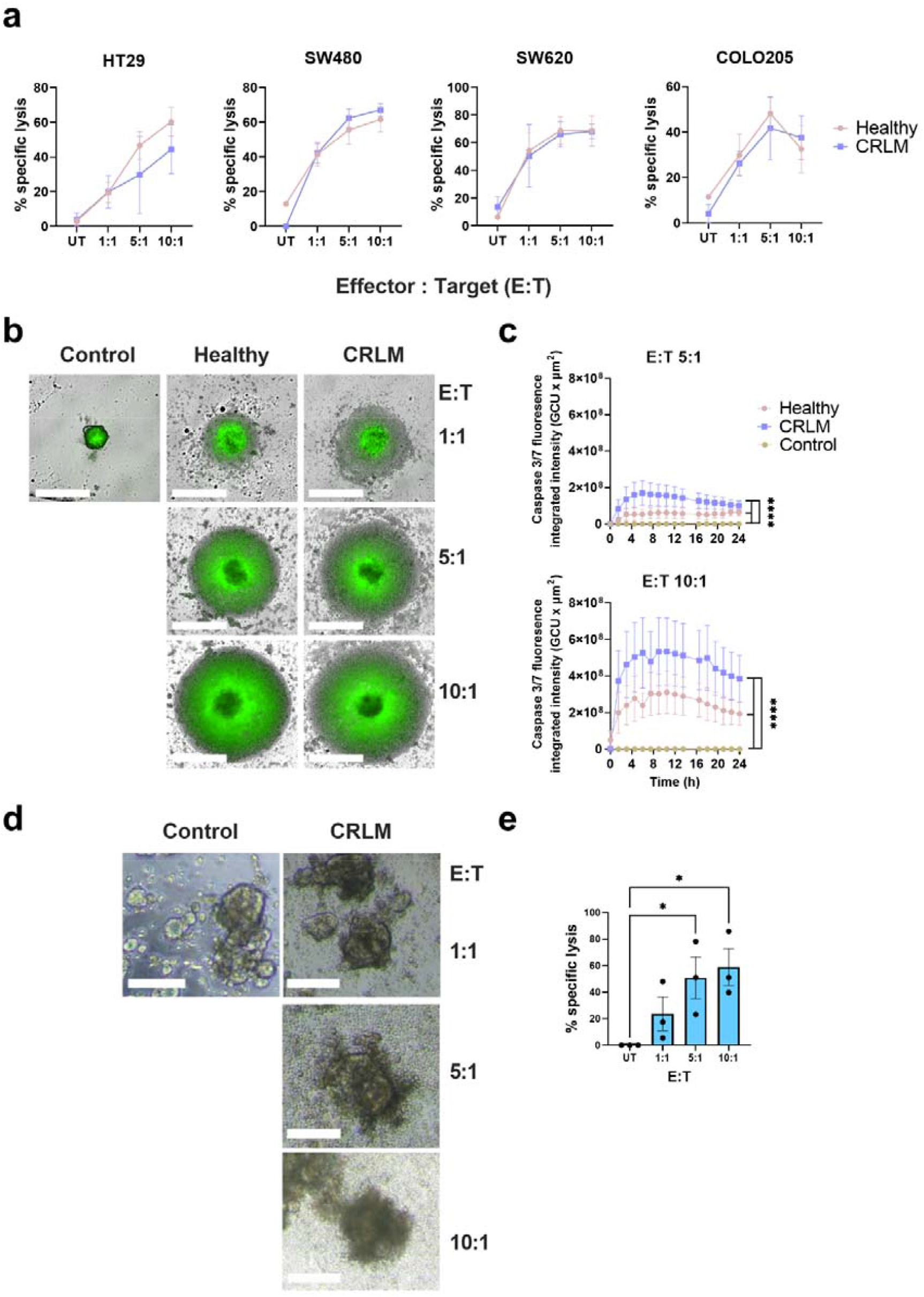
Cytotoxicity of CRLM donor-derived CIK cells. (a) 2D cytotoxicity assay with the HT29, COLO 205, SW480 or SW620 CRC cell lines as target cells and CIK cells at 10:1, 5:1, 1:1 effector to target (E:T) ratios. Target cells only were used as untreated (UT) controls. (b) 3D cytotoxicity assay with the HT-29 CRC cell line grown as spheroids as target cells and CIK cells at 10:1, 5:1, 1:1 E:T ratios, monitored over time using an Incucyte Live-Cell analysis system, (c) dynamic cleavage of caspase 3/7 (green) as a marker for cytotoxic activity measured over 24 hours. White bars represent 100μm. Caspase 3/7 graph shows mean ± SEM of triplicates. Data is representative of four independent experiments from separate donors. *\*\*\*\*p < 0*.*0005*. Two-way ANOVA with multiple comparison test were performed. (d) Autologous patient-derived tumour organoid (PDTO) 3D cytotoxicity assay (n=3). Representative images of CRLM PDTO (PID-0169) disruption after co-culture with autologous CIK cells at different E:T ratios for 24 hours. White bars represent 100μm. (e) The absolute number of live CellTrace™ Violet labelled PDTOs was obtained by flow cytometry and % specific lysis was calculated in duplicate or triplicate. Results shown are mean ± SEM from three independent experiments using a separate donor each time. *\*p* ≤ *0*.*05*. One-way ANOVA with multiple comparison test were performed.

### 3.3 Effect of patient characteristics on CIK cell production

Many studies have shown that biological sex has effects in cells of the adaptive immune response [31, 32]. We compared CIK cultures from females (n=3) and males (n=5) and found no significant difference in the number or percentage of CD3+CD56-, CD3+CD56+, CD3-CD56+ subpopulations on day 21 post-culture (Fig. 5a-b). Sex did not affect the percentage of CD8+CD3+CD56- or CD8+CD3+CD56+ cells (Fig. 5c) nor did it influence the expression of granzyme B or perforin (Fig. 5d) or the production of type-1 cytokines (Fig. 5e).

**Figure 5.**
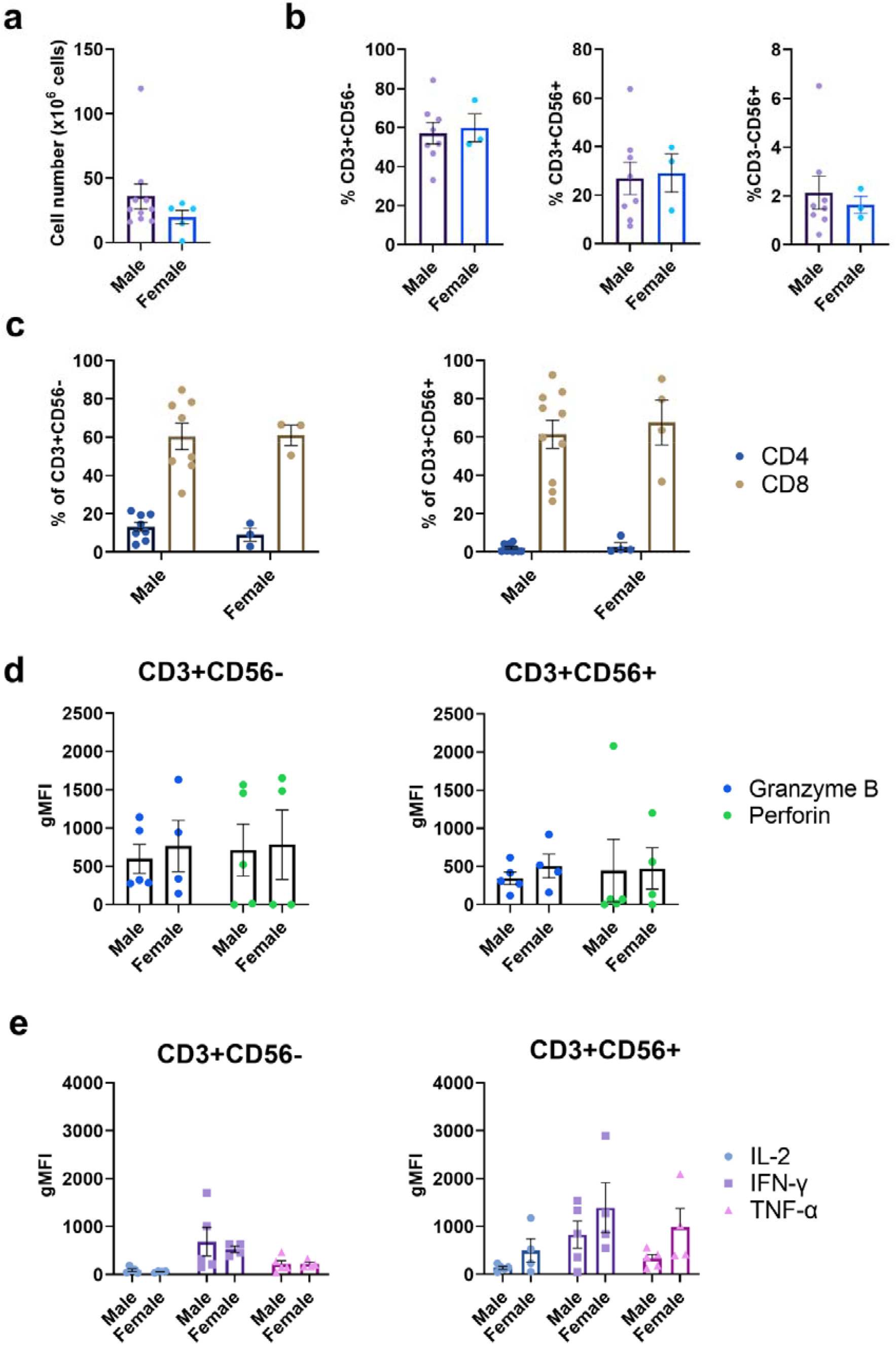
The effect of biological sex on CIK cells. (a) Total cell counts on day 21 post-culture for male and female healthy and CRLM donors. (b) Percentages of T (CD3+CD56-), NK-like T cells (CD3+CD56+) and NK cells (CD3-CD56+) in day 21 post-CIK cell culture. (c) The percentage of CD4+ and CD8+ subsets within the CD3+CD56- and CD3+CD56+ subpopulations at day 21 post-culture. (d) The gMFI for intracellular granzyme B and perforin and (e) IL-2, IFN-γ and TNF-α in CD8+CD3+CD56- and CD8+CD3+CD56+ CIK cells. Data are shown as mean ± SEM with each point representing an individual donor.

Most patients with cancer tend to be older, and the immune system tends to deteriorate with age [33]. Since the current preference is to use autologous PBMC to generate CIK cells for patient use, it is important to know if age can affect the number or quality of CIK cells generated. We compared CIK cells generated from younger (<55 years) and older (>55 years) donors with CRLM and younger (<55 years) healthy donors. We found no differences between these three groups in the total number of cells generated, the number or percentages of the subpopulations, or the expression of granzyme B or perforin, or the expression of the cytokines IL-2, IFN-γ or TNF-α (Fig. 6).

**Figure 6.**
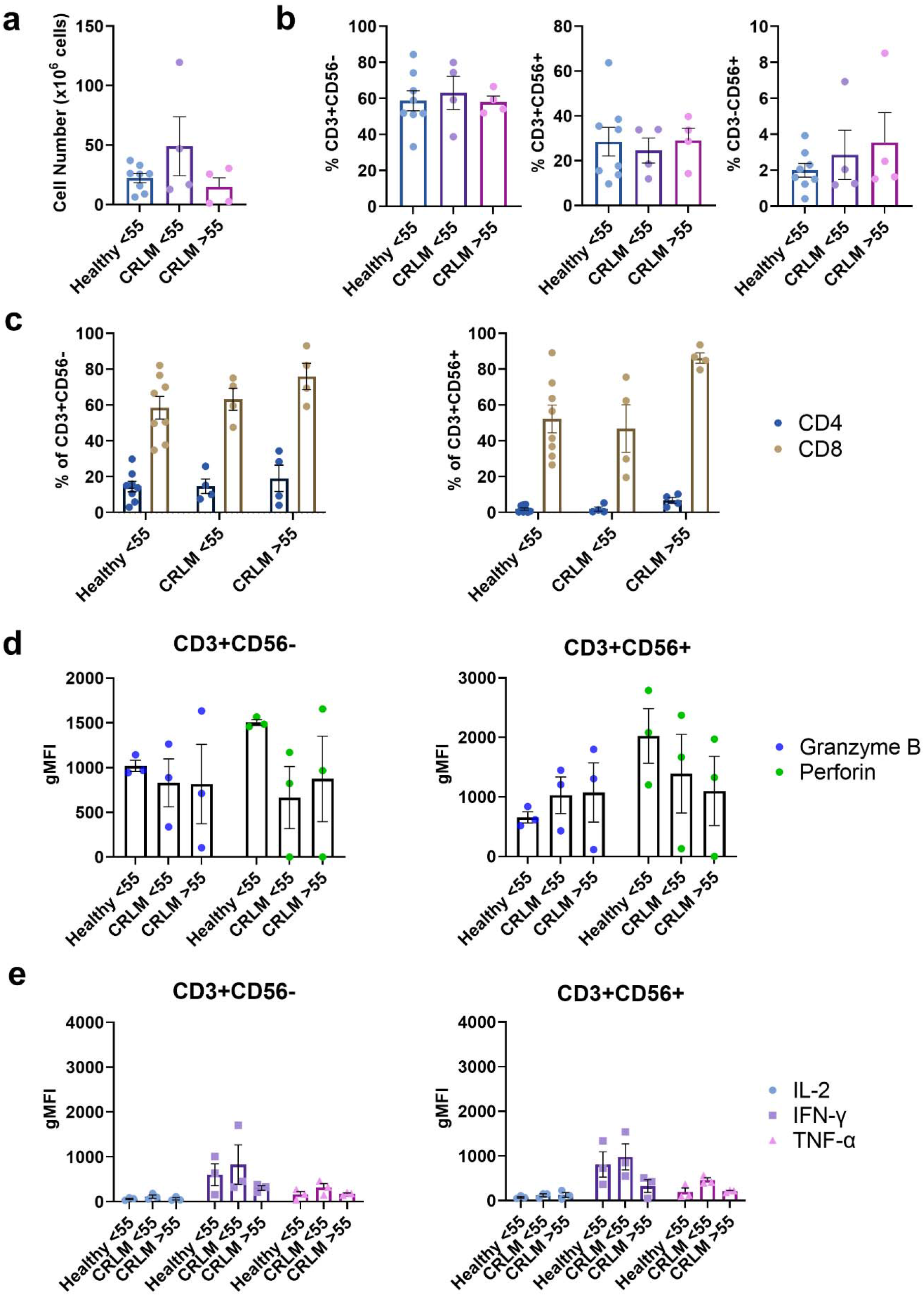
The effect of age on CIK cells. (a) Total cell counts on day 21 post-culture for different donor ages (Healthy <55 years, CRLM <55 years and CRLM >55 years). (b) Percentages of T (CD3+CD56-), NK-like T (CD3+CD56+) and NK cells (CD3-CD56+) at day 21 post-culture. (c) Percentage of CD4+ and CD8+ subsets within CD3+CD56- cells and CD3+CD56+ subpopulations at day 21 post-culture. (d) The gMFI intracellular granzyme B and perforin and (e) IL-2, IFN-γ, TNF-α in CD8+CD3+CD56- and CD8+CD3+CD56+ CIK cells. Data are shown as mean ± SEM with each point representing an individual donor.

It has been suggested that recent chemotherapy treatment can have long term effects on T cells that can impede the effectiveness of adoptive T cell therapies [34]. The impact of prior chemotherapy on autologous CIK cell generation is unknown. PBMCs from our patient cohort were collected just before curative-intent liver resection (Table 1). Importantly, a subset had received neoadjuvant chemotherapy prior to PBMC collection, which allowed us to address the influence of chemotherapy on CIK cell generation and function. We found no differences between chemotherapy naïve and treated patients in the total number of CIK cells generated, subset composition, or expression of the functional molecules granzyme B, perforin, IL-2, IFN-γ or TNF-α (Fig. 7). Taken together, we show that production of CIK cells is robust and highly feasible for a broad range of CRLM patients.

**Figure 7.**
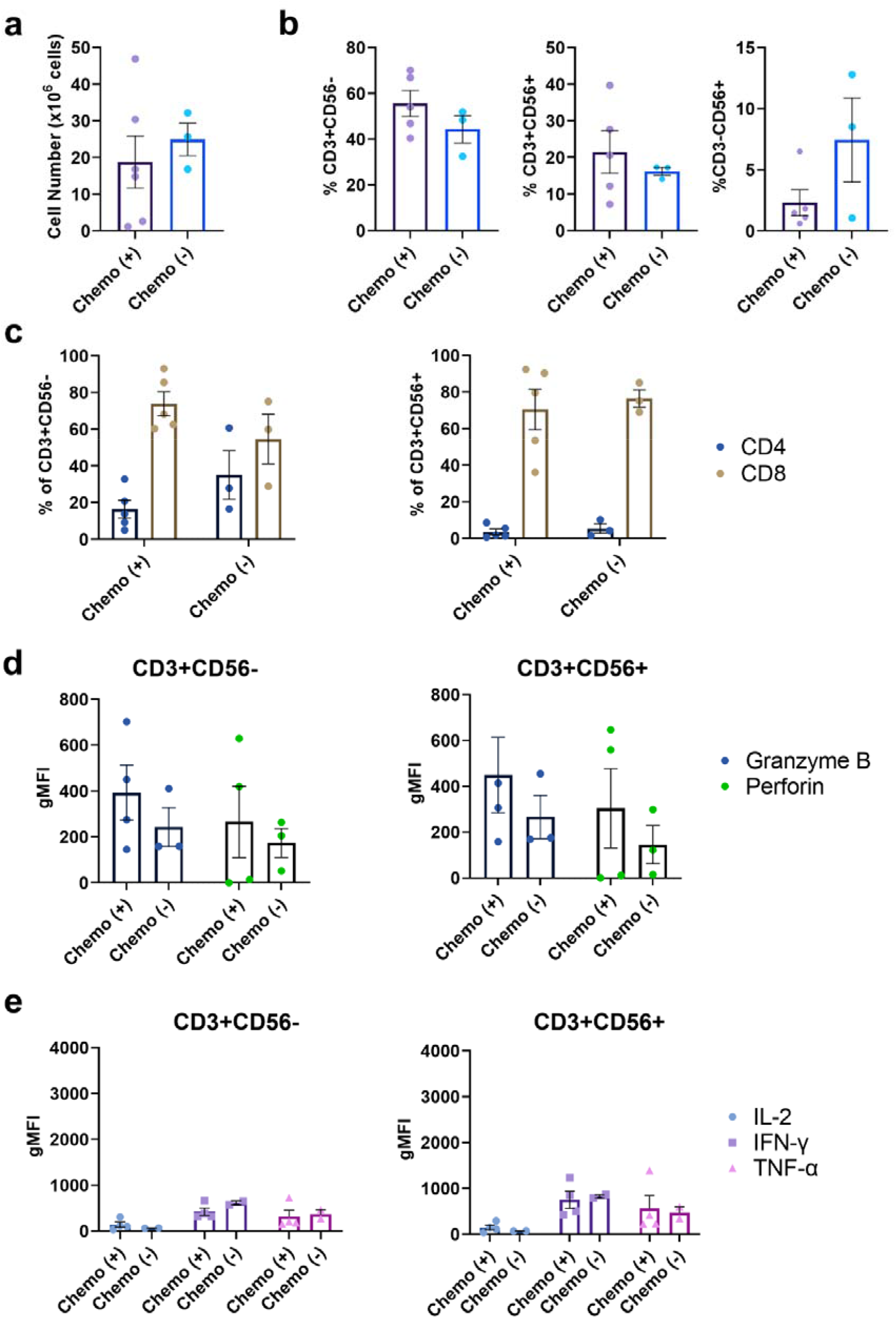
The effect of chemotherapy exposure on CIK cells. (a) Total cell counts on day 21 post-CIK cell culture for CRLM donors having received chemotherapy (+) or not (-) prior to PBMC collection. (b) Percentages of T (CD3+CD56-), NK-like T (CD3+CD56+) and NK cells (CD3-CD56+) at day 21 post-culture. (c) Percentage of CD4+ and CD8+ subsets within CD3+CD56- cells and CD3+CD56+ subpopulations at day 21 post-culture. (d) The gMFI intracellular granzyme B and perforin and (e) IL-2, IFN-γ, TNF-α in CD8+CD3+CD56- and CD8+CD3+CD56+ CIK cells. Data are shown as mean ± SEM with each point representing an individual CRLM donor.

## Discussion

Despite the introduction of a number of new management strategies for patients with CRLM, 5-year survival remains poor and new treatments are required.[35, 36]. Adoptive cell therapies, such as CIK cell therapy, are being actively investigated for treatment of a number of cancers. Reports that CIK cell therapy may result in significant clinical benefit in solid tumours including CRC suggests that wider adoption should be considered. However, in clinical studies the CIK cell culture methods can differ in detail and are often poorly described. For example, in four studies reporting CIK cell therapy for CRLM each used a different production protocol [14, 28, 37, 38]. Hence, this study aimed to develop a defined culture protocol for producing CIK cells for future CRLM clinical trials in Australia. We report that X-VIVO 15, a GMP grade SFM, in terms of CIK cell expansion, phenotypes, and cytotoxic capacities, was comparable to serum supplemented RPMI. We found no significant difference between CIK cells generated from healthy controls to those from patients with CRLM. Importantly, we determined that patient characteristics such as age, sex or prior chemotherapy exposure had minimal effect on CIK cell expansion and functionality.

Media usage ranged from SFMs to serum supplemented media including the use of human serum and FBS ([11] and unpublished observations). While FBS supplementation is still common practice in cell culture for T cell therapies [39], its use increases the risk of xenoimmunisation and zoonotic disease transfer in recipients [16]. Thus, we focused on comparing 3 SFMs specifically made for clinical grade T cell products. Media selected were based on their popularity in the adoptive T cell therapy field and accessibility in the Australian market. Interestingly, only with X-VIVO 15 were the CIK cells numbers comparable to complete RPMI. We observed only minor differences in the X-VIVO 15 cultured cells, including lower levels of DNAM-1 and cytokine expression. This may be overcome by modifying the culture method, such as the addition of IL-15 [40]. Importantly, X-VIVO 15 cultured CIK cells were comparably cytotoxic against CRC cells. Thus X-VIVO 15 can be used as the base medium for large-scale CIK cell production for clinical trials.

In our hands long-term storage (6-12 months) of cryopreserved CIK cells reduced their cytotoxic capacity. Our cryopreservation method used FBS supplemented with 10% DMSO and was done in a small scale to suit laboratory investigation. It has been previously reported that supplementation with IL-2 after thawing may rejuvenate CIK cell functionality [41]. Two recent reports utilising clinical scale CIK cell cryopreservation techniques showed that long-term storage of CIK cells has minimal effects on their functionality [42, 43].

T cell fitness, which is influenced by chronic infection, aging, cancer, and cancer treatment, is an emerging important factor in determining T cell manufacturing efficacy [44]. With our limited sample size, we found that age, sex and prior chemotherapy exposure did not influence CIK cell production in CRLM patients. Importantly, CRLM patients could generate the same numbers of CIK cells as healthy donors. Other factors such as infection history may have caused the poor expansion in a small number of CRLM and healthy donors, however we did not have access to complete patient data to address this. Currently, there is no consensus on how CIK cells are to be verified as functional prior to patient transfusion. This is the first report showing that autologous CIK cells are cytotoxic to matched PDTOs. Clinical trials with PDTO cytotoxicity assays as pre-screens are required to determine if this predicts efficacy *in vivo* [45].

In conclusion, this is the first study to show that clinical grade CIK cells can be successfully generated in an Australian cohort of patients with CRLM. The production of CIK cells using a generic serum free CIK cell production protocol appears robust, with most patient factors not affecting the numbers or function of the CIK cells produced. Together this supports further investigation on the usage of CIK cell therapy for the treatment of CRLM in Australia.

## Supporting information

Edited

## Abbreviations

Abbreviations: Definitions
CAPOX: Capecitabine and oxaliplatin
ChRT: Concurrent chemoradiotherapy
CO_2_: Carbon dioxide
CIK: Cytokine-induced killer
CRC: Colorectal cancer
CRLM: Colorectal cancer liver metastases
DMEM: Dulbecco’s Modified Eagle Medium
DMSO: Dimethyl sulfoxide
DNAM-1: DNAX Accessory Molecule-1
DPBS: Dulbecco’s phosphate buffered saline
EDTA: Ethylenediaminetetraacetic acid
FACS: Fluorescence-activated cell sorting
FASL: Fas ligand
FBS: Fetal bovine serum
FOLFOX: Folinic acid, fluorouracil and oxaliplatin
GMP: Good manufacturing practice
h-EGF: Human epidermal growth factor
HEPES: N-2-hydroxyethylpiperazine-N’-2-ethanesulfonic acid
IFN-γ: Interferon-gamma
IL-2: Interleukin-2
MHC: Major histocompatibility complex
N-acetyl-L: cyst N-acetyl-L-cysteine
NK: Natural killer
NKG2D: Natural-killer group 2 member D
PBMC: Peripheral blood mononuclear cell
PBS: Phosphate-buffered saline
PDTO: Patient-derived tumour organoid
Pen-Strep: Penicillin-streptomycin
Phenol-red: Phenolsulfonphthalein-red
RPMI: Roswell Park Memorial Institute
RT: Radiotherapy
SFM: Serum-free media
TNF-α: Tumour neurosis factor-alpha
TNT: Total-neoadjuvant therapy
TRAIL: TNF-related apoptosis-inducing ligand

## Author Contributions

Conceptualisation C.L. and K.F.; Collection and/or assembly of data: C.L., Y.T., B.D., R.,L., T.T., J.W., L.V., Data Analysis and Interpretation: C.L., Y.T., P.D., and K.F., resources: S.L.W., G.M., T.P., and K.F.; writing— original draft preparation, C.L., P.D., and K.F.; writing—review and editing, C.L.,P.D., E.S.,Y.T. and K.F.; supervision, G.M., T.P., P.D., funding acquisition, C.L., and K.F.; All authors have read and agreed to the published version of the manuscript.

## Data Availability

All data generated or analysed during this study are included in this published article and its supplementary information files.

## Ethics Approval

This study was approved by the Human Research Ethics Committee of the Central Adelaide Local Health Network under protocol number HREC/14/TQEHLMH/164.

## Consent to participate

Informed consent was obtained from all individual participants included in the study.

## Reference

[1] “The Global Cancer Observatory 2020; Colorectal Cancer,” 2020, doi: https://gco.iarc.fr/today/data/factsheets/cancers/10_8_9-Colorectum-fact-sheet.pdf.

[2] H. Zhou et al., “Colorectal liver metastasis: molecular mechanism and interventional therapy,” Signal Transduction and Targeted Therapy, vol. 7, no. 1, p. 70, 2022/03/04 2022, doi: 10.1038/s41392-022-00922-2.

[3] K. Menck et al., “High-Throughput Profiling of Colorectal Cancer Liver Metastases Reveals Intra- and Inter-Patient Heterogeneity in the EGFR and WNT Pathways Associated with Clinical Outcome,” (in eng), Cancers, vol. 14, no. 9, Apr 21 2022, doi: 10.3390/cancers14092084.

[4] J. Martin et al., “Colorectal liver metastases: Current management and future perspectives,” (in eng), World J Clin Oncol, vol. 11, no. 10, pp. 761–808, Oct 24 2020, doi: 10.5306/wjco.v11.i10.761.

[5] X. Li et al., “Retrospective analysis of the efficacy of adjuvant cytokine-induced killer cell immunotherapy combined with chemotherapy in colorectal cancer patients after surgery,” (in eng), Clinical & translational immunology, vol. 11, no. 1, p. e1368, 2022, doi: 10.1002/cti2.1368.

[6] M. Riihimäki, A. Hemminki, J. Sundquist, and K. Hemminki, “Patterns of metastasis in colon and rectal cancer,” (in eng), Scientific reports, vol. 6, p. 29765, Jul 15 2016, doi: 10.1038/srep29765.

[7] Y. Guo and W. Han, “Cytokine-induced killer (CIK) cells: from basic research to clinical translation,” (in eng), Chinese journal of cancer, vol. 34, no. 3, pp. 99–107, Mar 5 2015, doi: 10.1186/s40880-015-0002-1.

[8] J. Jiang, C. Wu, and B. Lu, “Cytokine-induced killer cells promote antitumor immunity,” (in eng), Journal of translational medicine, vol. 11, p. 83, Mar 28 2013, doi: 10.1186/1479-5876-11-83.

[9] X. Gao et al., “Cytokine-Induced Killer Cells As Pharmacological Tools for Cancer Immunotherapy,” (in eng), Frontiers in immunology, vol. 8, p. 774, 2017, doi: 10.3389/fimmu.2017.00774.

[10] L. Zhang et al., “Cytokine-induced killer cells/dendritic cells-cytokine induced killer cells immunotherapy combined with chemotherapy for treatment of colorectal cancer in China: a metaanalysis of 29 trials involving 2,610 patients,” (in eng), Oncotarget, vol. 8, no. 28, pp. 45164–45177, Jul 11 2017, doi: 10.18632/oncotarget.16665.

[11] C. M. Ying Li et al., “Use of cytokine-induced killer cell therapy in colorectal cancer patients: a systematic review and meta-analysis,” medRxiv, p. 2023.01.05.22283441, 2023, doi: 10.1101/2023.01.05.22283441.

[12] Wang R, Meng M L. R. Li P, Zhao W, and H. Z, “Effects of IL-2- and IL-15-induced CIK Cells Combined with Chemotherapy Treatment for Colorectal Cancer (Chinese Article),” Journal of Junming Medical University vol. 35, no. 11, pp. 97–101, 2014, doi: 10.3969/j.issn.1003-4706.2014.11.025.

[13] C. Du, Z. Liu, Z. Ding, F. Guo, D. Ma, and X. Xie, “Autologous cytokine-induced killer cells combined with chemotherapy in the treatment of advanced colorectal cancer: a randomized control study,” The Chinese-German Journal of Clinical Oncology, vol. 12, no. 10, pp. 487–491, 2013/10/01 2013, doi: 10.1007/s10330-013-1214-y.

[14] X. Li et al., “Phase II/III Study of Radiofrequency Ablation Combined with Cytokine-Induced Killer Cells Treating Colorectal Liver Metastases,” (in eng), Cellular physiology and biochemistry : international journal of experimental cellular physiology, biochemistry, and pharmacology, vol. 40, no. 1-2, pp. 137–145, 2016, doi: 10.1159/000452531.

[15] Li Y, Jin A, Chen S, Song C, and Z. G., “Efficacy of Adjuvant Chemotherapy Combined with CIK Cell Immunotherapy in 130 Patients with Postoperative Colorectal Cancer (Chinese Article),” Journal of Chinese Oncology, vol. 21, no. 10, pp. 843–847, 2015, doi: 10.11735/j.issn.1671-170X.2015.10.B013.

[16] A. Mackensen, R. Dräger, M. Schlesier, R. Mertelsmann, and A. Lindemann, “Presence of IgE antibodies to bovine serum albumin in a patient developing anaphylaxis after vaccination with human peptide-pulsed dendritic cells,” (in eng), Cancer immunology, immunotherapy : CII, vol. 49, no. 3, pp. 152–6, Jun 2000, doi: 10.1007/s002620050614.

[17] H. W. Grievink, T. Luisman, C. Kluft, M. Moerland, and K. E. Malone, “Comparison of Three Isolation Techniques for Human Peripheral Blood Mononuclear Cells: Cell Recovery and Viability, Population Composition, and Cell Functionality,” (in eng), Biopreserv Biobank, vol. 14, no. 5, pp. 410–415, Oct 2016, doi: 10.1089/bio.2015.0104.

[18] V. Narasimhan et al., “Medium-throughput Drug Screening of Patient-derived Organoids from Colorectal Peritoneal Metastases to Direct Personalized Therapy,” (in eng), Clinical cancer research : an official journal of the American Association for Cancer Research, vol. 26, no. 14, pp. 3662–3670, Jul 15 2020, doi: 10.1158/1078-0432.Ccr-20-0073.

[19] C. M. Cattaneo et al., “Tumor organoid-T-cell coculture systems,” (in eng), Nat Protoc, vol. 15, no. 1, pp. 15–39, Jan 2020, doi: 10.1038/s41596-019-0232-9.

[20] X. Wu, Y. Zhang, Y. Li, and I. G. H. Schmidt-Wolf, “Improvements in Flow Cytometry-Based Cytotoxicity Assay,” (in eng), Cytometry A, vol. 99, no. 7, pp. 680–688, Jul 2021, doi: 10.1002/cyto.a.24242.

[21] “Incucyte® Immune Cell Killing of Tumour Spheroids Assay.” https://www.sartorius.com/download/829112/incucyte-immune-cell-killing-tumor-spheroids-assay-protocol-1--data.pdf x(accessed 31/10/2022.

[22] S. Liu et al., “CD4(+) T cells are required to improve the efficacy of CIK therapy in non-small cell lung cancer,” (in eng), Cell death & disease, vol. 13, no. 5, p. 441, May 6 2022, doi: 10.1038/s41419-022-04882-x.

[23] D. Sangiolo, “Cytokine induced killer cells as promising immunotherapy for solid tumors,” (in eng), Journal of Cancer, vol. 2, pp. 363–8, 2011, doi: 10.7150/jca.2.363.

[24] I. Voskoboinik, J. C. Whisstock, and J. A. Trapani, “Perforin and granzymes: function, dysfunction and human pathology,” Nature Reviews Immunology, vol. 15, no. 6, pp. 388–400, 2015/06/01 2015, doi: 10.1038/nri3839.

[25] A. Pievani et al., “Dual-functional capability of CD3+CD56+ CIK cells, a T-cell subset that acquires NK function and retains TCR-mediated specific cytotoxicity,” (in eng), Blood, vol. 118, no. 12, pp. 3301–10, Sep 22 2011, doi: 10.1182/blood-2011-02-336321.

[26] Y. C. Linn, S. M. Wang, and K. M. Hui, “Comparative gene expression profiling of cytokine-induced killer cells in response to acute myloid leukemic and acute lymphoblastic leukemic stimulators using oligonucleotide arrays,” Experimental hematology, vol. 33, no. 6, pp. 671–681, 2005/06/01/2005, doi: 10.1016/j.exphem.2005.03.005.

[27] H. T. Ngo, V. T. Dang, N. H.-T. Nguyen, A. N.-T. Bui, and P. Van Pham, “Comparison of cytotoxic potency between freshly cultured and freshly thawed cytokine-induced killer cells from human umbilical cord blood,” Cell and Tissue Banking, vol. 24, no. 1, pp. 139–152, 2023/03/01 2023, doi: 10.1007/s10561-022-10022-8.

[28] Q. Z. Pan et al., “Retrospective analysis of the efficacy of cytokine-induced killer cell immunotherapy combined with first-line chemotherapy in patients with metastatic colorectal cancer,” (in eng), Clinical & translational immunology, vol. 9, no. 2, p. e1113, 2020, doi: 10.1002/cti2.1113.

[29] W. Wu, X. Li, and S. Yu, “Patient-derived Tumour Organoids: A Bridge between Cancer Biology and Personalised Therapy,” Acta Biomaterialia, vol. 146, pp. 23–36, 2022/07/01/ 2022, doi: 10.1016/j.actbio.2022.04.050.

[30] J. Wang et al., “Patient-Derived Tumor Organoids: New Progress and Opportunities to Facilitate Precision Cancer Immunotherapy,” (in eng), Front Oncol, vol. 12, p. 872531, 2022, doi: 10.3389/fonc.2022.872531.

[31] S. L. Klein and K. L. Flanagan, “Sex differences in immune responses,” Nature Reviews Immunology, vol. 16, no. 10, pp. 626–638, 2016/10/01 2016, doi: 10.1038/nri.2016.90.

[32] I. Miguel-Aliaga, “Let’s talk about (biological) sex,” Nature Reviews Molecular Cell Biology, vol. 23, no. 4, pp. 227–228, 2022/04/01 2022, doi: 10.1038/s41580-022-00467-w.

[33] A. D. Foster, A. Sivarapatna, and R. E. Gress, “The aging immune system and its relationship with cancer,” (in eng), Aging health, vol. 7, no. 5, pp. 707–718, Oct 1 2011, doi: 10.2217/ahe.11.56.

[34] R. K. Das, R. S. O’Connor, S. A. Grupp, and D. M. Barrett, “Lingering effects of chemotherapy on mature T cells impair proliferation,” (in eng), Blood Adv, vol. 4, no. 19, pp. 4653–4664, Oct 13 2020, doi: 10.1182/bloodadvances.2020001797.

[35] R. Adam and Y. Kitano, “Multidisciplinary approach of liver metastases from colorectal cancer,” (in eng), Annals of gastroenterological surgery, vol. 3, no. 1, pp. 50–56, Jan 2019, doi: 10.1002/ags3.12227.

[36] Y. Lv et al., “Benefits of multi-disciplinary treatment strategy on survival of patients with colorectal cancer liver metastasis,” (in eng), Clin Transl Med, vol. 10, no. 3, p. e121, Jul 2020, doi: 10.1002/ctm2.121.

[37] H. Zhu et al., “Immune response, safety, and survival and quality of life outcomes for advanced colorectal cancer patients treated with dendritic cell vaccine and cytokine-induced killer cell therapy,” (in eng), BioMed research international, vol. 2014, p. 603871, 2014, doi: 10.1155/2014/603871.

[38] Y. Xie, L. Huang, L. Chen, X. Lin, L. Chen, and Q. Zheng, “Effect of dendritic cell-cytokine-induced killer cells in patients with advanced colorectal cancer combined with first-line treatment,” (in eng), World journal of surgical oncology, vol. 15, no. 1, p. 209, Nov 28 2017, doi: 10.1186/s12957-017-1278-1.

[39] N. Watanabe, F. Mo, and M. K. McKenna, “Impact of Manufacturing Procedures on CAR T Cell Functionality,” (in eng), Frontiers in immunology, vol. 13, p. 876339, 2022, doi: 10.3389/fimmu.2022.876339.

[40] C. Wei et al., “The CIK cells stimulated with combination of IL-2 and IL-15 provide an improved cytotoxic capacity against human lung adenocarcinoma,” (in eng), Tumour biology : the journal of the International Society for Oncodevelopmental Biology and Medicine, vol. 35, no. 3, pp. 1997–2007, Mar 2014, doi: 10.1007/s13277-013-1265-2.

[41] J. Baker, K. Sheehan, G. Monterola, N. Staines, and R. S. Negrin, “Human CIK Maintain Their In Vitro and In Vivo Anti-Tumor Ability after Cryopreservation,” Blood, vol. 106, no. 11, pp. 1062–1062, 2005, doi: 10.1182/blood.V106.11.1062.1062.

[42] M. Bremm et al., “Improving Clinical Manufacturing of IL-15 Activated Cytokine-Induced Killer (CIK) Cells,” (in eng), Frontiers in immunology, vol. 10, p. 1218, 2019, doi: 10.3389/fimmu.2019.01218.

[43] K. Mareschi et al., “Cytokine-Induced Killer (CIK) Cells, In Vitro Expanded under Good Manufacturing Process (GMP) Conditions, Remain Stable over Time after Cryopreservation,” (in eng), Pharmaceuticals (Basel), vol. 13, no. 5, May 12 2020, doi: 10.3390/ph13050093.

[44] P. H. Mehta, S. Fiorenza, R. M. Koldej, A. Jaworowski, D. S. Ritchie, and K. M. Quinn, “T Cell Fitness and Autologous CAR T Cell Therapy in Haematologic Malignancy,” (in eng), Frontiers in immunology, vol. 12, p. 780442, 2021, doi: 10.3389/fimmu.2021.780442.

[45] J. C. H. Kong et al., “Tumor-Infiltrating Lymphocyte Function Predicts Response to Neoadjuvant Chemoradiotherapy in Locally Advanced Rectal Cancer,” (in eng), JCO Precis Oncol, vol. 2, pp. 1-15, Nov 2018, doi: 10.1200/po.18.00075.

