## Supplementary material for "Generation and assessment of cytokine-induced killer cells for the treatment of colorectal cancer liver metastases": Edited

**Supplementary table 1.** Characteristics of patients with colorectal cancer liver metastases.

| Patient ID. | Sex | Age Range | Cancer origins | Timeline of conventional therapy received (year of treatment) | | | | Conventional therapy received before hepatectomy |
| --- | --- | --- | --- | --- | --- | --- | --- | --- |
|  |  |  |  | **1^st^ line** | **2^nd^ line** | **3^rd^ line** | **4^th^ line** |  |
| PID-0192 | F | 61-65 | Caecum | Hemicolectomy (2018) | FOLFOX complicated by port thrombosis | 12 cycles Irinotecan (2019-2020) | Right hepatectomy  (2020) | Y |
| PID-0193 | M | 76-80 | Rectum | Ch-5FU-RT | Hepatectomy (2019) | - | - | Y |
| PID-0195 | F | 46-50 | Splenic flexure | Left hemicolectomy (2020) | Right hepatectomy (2020) | - | - | N |
| PID-0196 (a) | M | 31-35 | Colon | 12 cycles of FOLFOX/panitumumab (2020) | Hepatectomy (2020) |  |  | Y |
| PID-0197 | M | 51-55 | Rectum | 2 cycles of FOLFOX (2019) | 4 cycles of CAPOX (2020) | 5 weeks of CAPOX-RT (2020) | 7 cycles of Irinotecan-panitumumab (2020),  liver surgery abandoned | Y |
| PID-0196 (b) | M | 31-35 | Colon | Hepatectomy for liver recurrence (2021) | - | - | - | N |
| PID-0198 | F | 61-65 | Rectum | TNT (FOLFOX or CAPOX, then ChRT) (2020) | Partial right hepatectomy (2021) | - | - | Y |
| PID-0190 | M | 46-50 | Caecum | Right hemicolectomy (2019) | 6 cycles of FOLFOX (2019-2020) | Hepatectomy (2020) | - | Y |
| PID-0199 | F | 46-50 | Sigmoid | High anterior resection (2021) | Hepatectomy (2021) | - | - | N |
| PID-0196 (c) | M | 30-35 | Colon | Hepatectomy for liver recurrence (2021) | - | - | - | N |
| PID-0191 | M | 66-70 | Colon | Anterior resection (2018) | Adjuvant CAPOX | Right hepatectomy for liver recurrence (2020) | - | Y |
| PID-0169 | M | 61-65 | Colon | Low anterior resection (2019) | Right hepatectomy (2021) liver recurrence | - | - | N |
| PID-0200 | M | 66-70 | Colon | High anterior resection (2020) | Hepatectomy (2021) for liver recurrence | - | - | N |
| PID-0201 (a) | M | 56-60 | Right colon | Right hemicolectomy (2020) | Adjuvant FOLFOX | Left hemi-hepatectomy for liver recurrence (2021) | - | Y |
| PID-0202 | M | 86-90 | Rectum | - | Left hemi-hepatectomy for liver recurrence (2021) | - | - | N |
| PID-0203 | M | 81-90 | Colon | ChRT (2019) | - | 4 cycles of Carboplatin-etoposide for recurrence in liver | Hepatectomy in (March 2022 and November 2022) | Y |
| PID-0204 | M | 56-60 | Caecum | Right hemicolectomy (2021) | Liver surgery abandoned (2022) | - | - | N |
| PID-0205 | M | 31-35 | Colon | 8 cycles of FOLFOX-panitumumab (2022) | Short course RT to rectum (2022) | Right hepatectomy (2022) | - | Y |
| PID-0201 (b) | M | 56-60 | Right colon | Left hemi-hepatectomy for liver recurrence (2022) | - | - | - | N |
| PID-0208 | M | 76-80 | Colon | Neoadjuvant chemotherapy (2022) | Neoadjuvant chemotherapy completed approx. 7 weeks ago (2022) | Hepatectomy (2022) | Hepatectomy for liver recurrence (2023), liver surgery abandoned | Y |

RT, Radiotherapy; TNT, Total neoadjuvant therapy; ChRT, Concurrent chemoradiotherapy; CAPOX, Capecitabine and oxaliplatin, PID-0196 (a-c) were the same patient; PID-0201 (a, b) were the same patient; Y, Yes; N, No.

**Supplementary table 2.** Characteristics of healthy donors.

| Donor ID. | Sex | Age Range |
| --- | --- | --- |
| 1 | M | (46-50) |
| 2 | M | (26-30) |
| 3 | M | (56-60) |
| 4 | M | (31-35) |
| 5 | F | (36-40) |
| 6 | F | (31-35) |
| 7 | M | (26-30) |
| 8 | M | (36-40) |

**Supplementary Table 3.** Statistical analyses of the absolute numbers in CIK cell subpopulations at day 21 in different culture media (Two-way ANOVA).

| **RPMI at day 21** | **#CD3+CD56-** | **#CD3+CD56+** | **#CD3-CD56+** |
| --- | --- | --- | --- |
| Mean ± SD | 14.96 ± 9.63 | 8.61 ± 5.99 | 2.47 ± 2.49 |
| Median (Range) | 13.365 (2.13-28.2) | 6.93 (1.51-18.83) | 1.33 (0.02-6.49) |
| **X-VIVO 15 at day 21** | **#CD3+CD56-** | **#CD3+CD56+** | **#CD3-CD56+** |
| Mean ± SD | 13.27 ± 7.41 | 5.93 ± 3.94 | 0.44 ± 0.31 |
| Median (Range) | 13.3641 (2.08-27.42) | 4.5362725 (0.86-12.7) | 0.5 (0.08-1.03) |
| **TexMACS at day 21** | **#CD3+CD56-** | **#CD3+CD56+** | **#CD3-CD56+** |
| Mean ± SD | 1.3945828 ± 0.85 | 0.1693914 ± 0.11 | 0.04849 ± 0.03 |
| Median (Range) | 1.64985 (0.04-0.32) | 0.14076 (0.16-2.37) | 0.04692 (0-0.09) |
| **CTS OpTmizer at day 21** | **#CD3+CD56-** | **#CD3+CD56+** | **#CD3-CD56+** |
| Mean ± SD | 0.18180475 ± 0.23 | 0.0205553375 ± 0.027 | 0.128797 ± 0.18 |
| Median (Range) | 0.099285 (0-0.06) | 0.01123275 (0-0.52) | 0.058065 (0-0.4) |

| ***p values*** | **#CD3+CD56-** | **#CD3+CD56+** | **#CD3-CD56+** |
| --- | --- | --- | --- |
| **RPMI vs X-VIVO 15** | 0.9427 (ns) | 0.3133 (ns) | *<0.0001* |
| **RPMI vs TexMACS** | *<0.0001* | *<0.0001* | *<0.0001* |
| **X-VIVO 15 vs TexMACS** | *<0.0001* | *0.0004* | 0.7643 (ns) |
| **RPMI vs CTS OpTmizer** | *<0.0001* | *<0.0001* | *<0.0001* |
| **X-VIVO 15 vs CTS OpTmizer** | *<0.0001* | *0.0008* | 0.8885 (ns) |
| **CTS OpTmier vs TexMACS** | 0.9705 (ns) | 0.9997 (ns) | 0.9982 (ns) |

**Supplementary Table 4.** Statistical analyses of the percentages of CD8+ and CD4+ cells in CIK cell subpopulations at day 21 in RPMI and X-VIVO 15 media (Two-way ANOVA).

| Day 21 | %CD8+CD3+CD56- | %CD8+CD3+CD56+ | %CD4+CD3+CD56- | %CD4+CD3+CD56+ |
| --- | --- | --- | --- | --- |
| **RPMI** |  |  |  |  |
| Mean ± SD | 47.39 ± 21.70 | 71.53 ± 17.19 | 12.79 ± 6.62 | 0.98 ± 0.94 |
| Median (Range) | 48.7 (16.6-71.4) | 71.4 (43.3-88.7) | 14.3 (2.75-21) | 0.82 (0.17-2.76) |
| **X-VIVO 15** |  |  |  |  |
| Mean ± SD | 53.27 ± 25.82 | 51.84 ± 21.91 | 18.17 ± 14.58 | 1.77 ± 1.91 |
| Median (Range) | 58.5 (9.97-78.4) | 48.95 (26.5-89.1) | 14.8 (2.98-47.5) | 0.905 (0.16-4.47) |
| **RPMI vs X-VIVO 15** *p value* | 0.9828 (ns) | 0.2745 (ns) | 0.8113 (ns) | 0.993 (ns) |

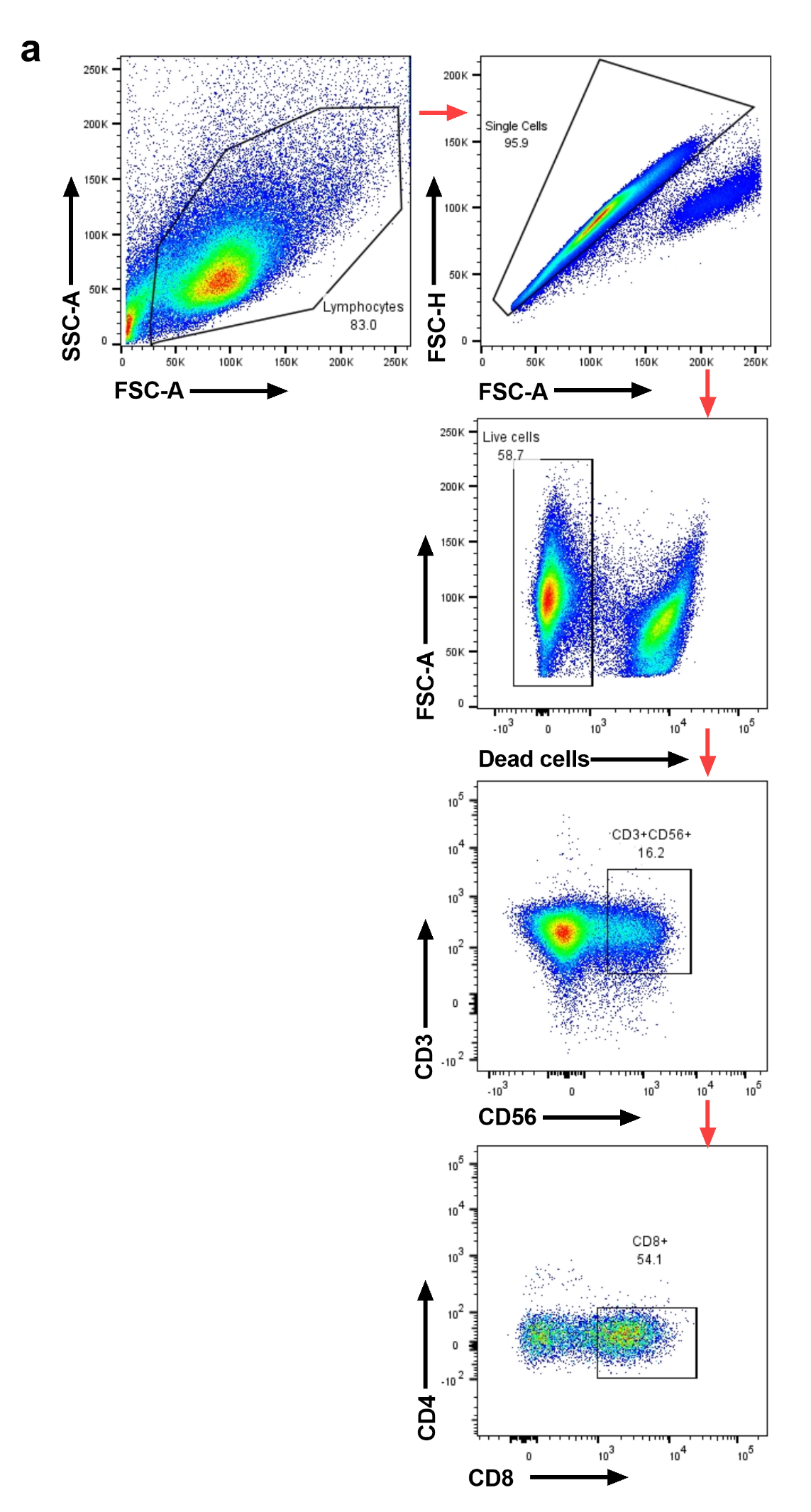

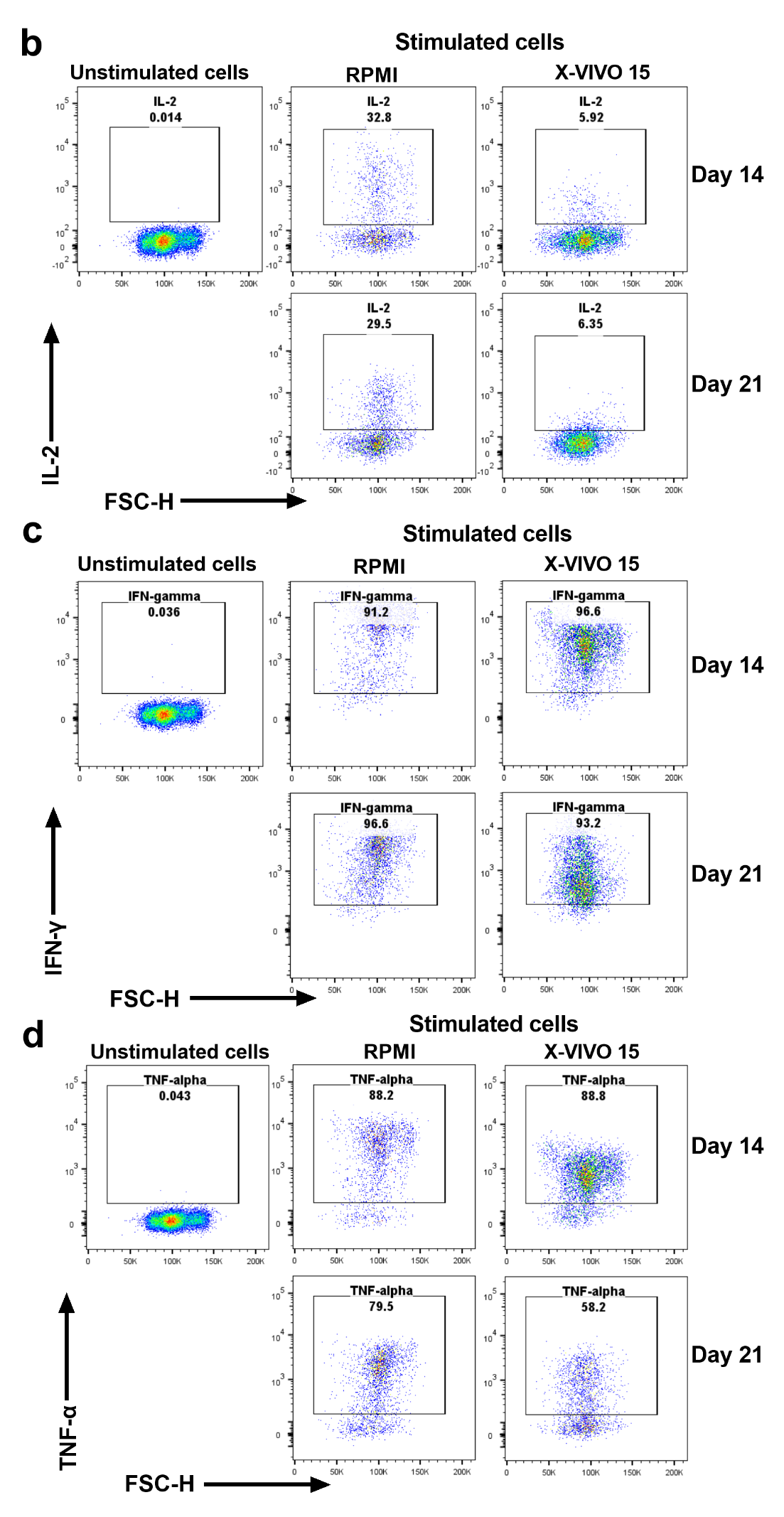

**Supplementary Figure 1.** Cytokine expression in CD8+CD3+CD56+ subpopulations of CIK cells generated in RPMI or X-VIVO 15. (a) Representative FACS plots showing the pre-gating strategy (see arrows) for cytokine expression in lymphocytes, single cells, live cells, CD3+CD56+ and CD8+ cells. (b) IL-2, (c) IFN-γ and (d) TNF-α expression in unstimulated cells and cells stimulated with Cytokine Activation Cocktail generated in RPMI or X-VIVO 15 at days 14 and 21 post-culture.

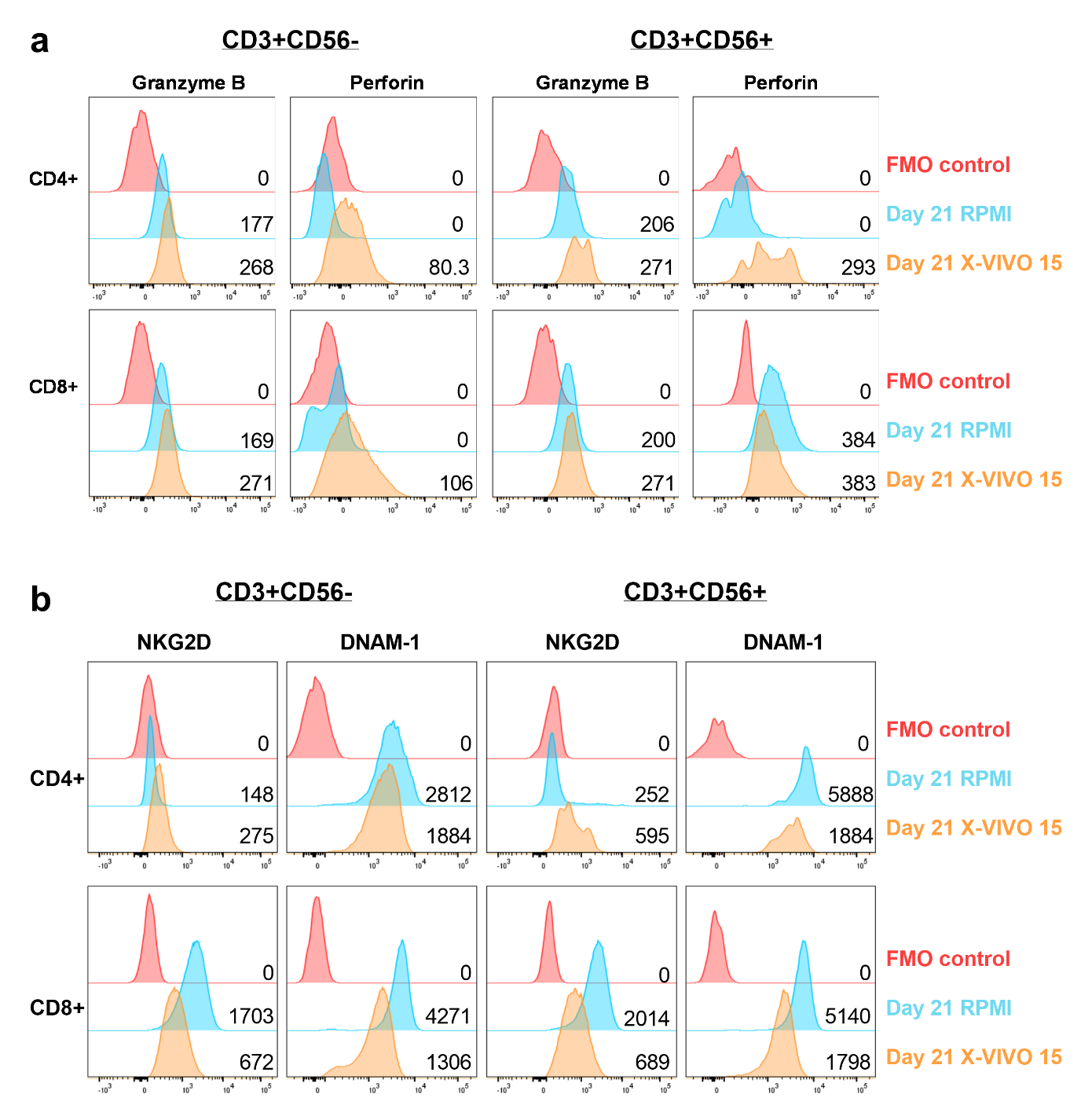

**Supplementary Figure 2.** The expression of activating receptors and intracellular cytotoxic molecules in CD4+ and CD8+ subsets of the CD3+CD56- and CD3+CD56+ subpopulations of CIK cells generated in RPMI or X-VIVO 15 at day 0 and day 21 post-culture. (a) NKG2D and DNAM-1 expression. (b) Granzyme B and Perforin expression.

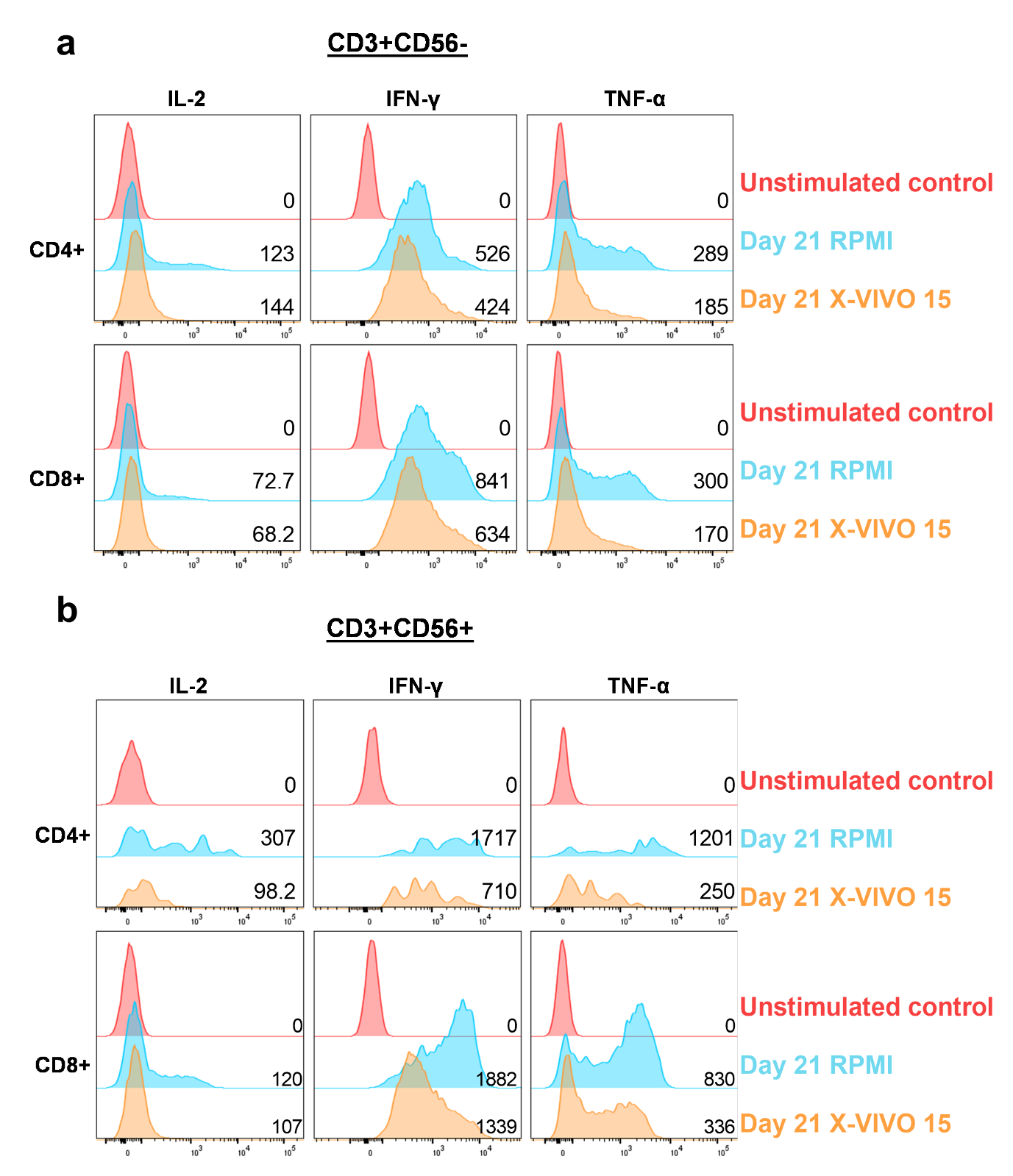
**Supplementary Figure 3.** The expression of cytokines in CD4+ and CD8+ subsets from CD3+CD56- and CD3+CD56+ subpopulations of CIK cells generated in RPMI or X-VIVO 15 at day 21 post-culture. (a) IL-2, IFN-γ, TNF-α expression in the CD3+CD56- subpopulations. (b) IL-2, IFN-γ, TNF-α expression in the CD3+CD56+ subpopulations.

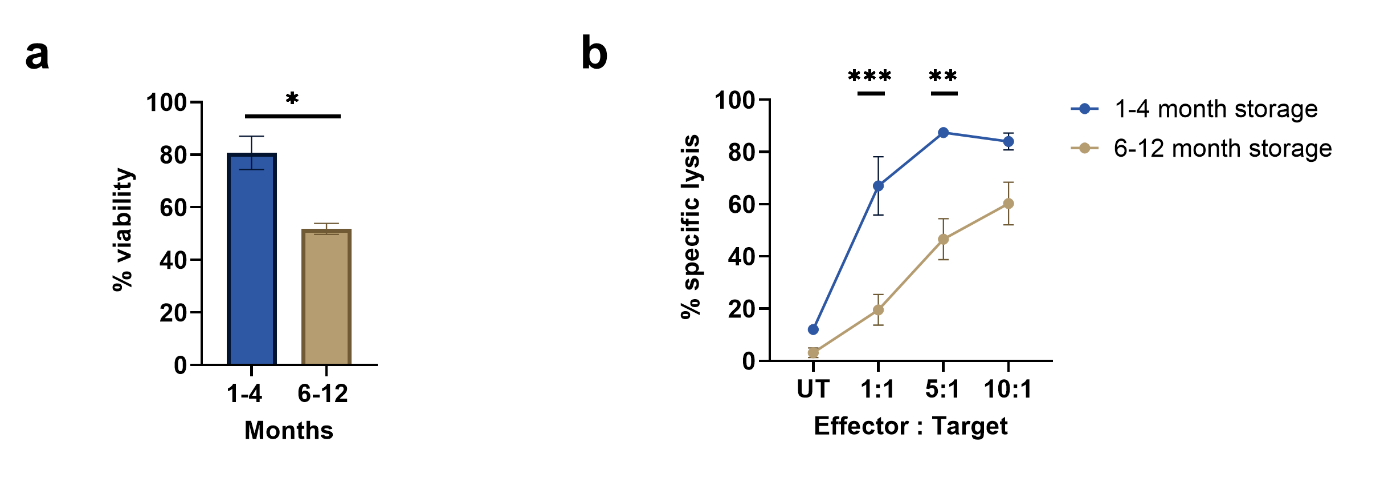

**Supplementary Figure 4.** The effect of long-term cryopreservation on CIK cells. (**a**) Viability of thawed CIK cells and (**b**) percentage of specific lysis against HT-29 monolayers using CIK therapy products stored between 1-4 or 6-12 months (n=3). **p ≤ 0.05, ** p ≤ 0.01, *** p ≤ 0.005*. Unpaired t test was performed to analyse the % of viability and two-way ANOVA with multiple comparison test for % of specific lysis.
